# A Multiple Session Dataset of NIRS Recordings From Stroke Patients Controlling Brain–Computer Interface

**DOI:** 10.1101/2024.03.27.24304842

**Authors:** Mikhail R. Isaev, Olesya A. Mokienko, Roman Kh. Lyukmanov, Ekaterina S. Ikonnikova, Anastasiya N. Cherkasova, Nataliya A. Suponeva, Michael A. Piradov, Pavel D. Bobrov

## Abstract

This paper presents an open dataset of over 50 hours of near infrared spectroscopy (NIRS) recordings. Fifteen stroke patients completed a total of 237 motor imagery brain–computer interface (BCI) sessions. The BCI was controlled by imagined hand movements; visual feedback was presented based on the real– time data classification results. We provide the experimental records, patient demographic profiles, clinical scores (including ARAT and Fugl–Meyer), online BCI performance, and a simple analysis of hemodynamic response. We assume that this dataset can be useful for evaluating the effectiveness of various near– infrared spectroscopy signal processing and analysis techniques in patients with cerebrovascular accidents.

## Background & Summary

Brain**–**computer interfaces (BCIs) provide a technological solution to convert data on the brain electrical or metabolic activity into control signals for an external device. BCIs can be used to provide feedback during motor imagery training (i.e. ideomotor training), which is one of the methods for motor rehabilitation after stroke [1,2]. Between 2019 and 2023, at least 11 systematic reviews were published, 8 of which included meta–analyses, demonstrating the efficacy of post**–**stroke BCI training [3–13]. It is important to note that there is a target group of patients for BCI training: those with severe paresis in the early stages after a stroke who are unable to partake in traditional physical therapy [2].

BCI technology that registers the electroencephalographic (EEG) signal accompanying the motor imagery process is the most studied for clinical application. However, EEG**–**BCI might not be practical for routine clinical use, due to high sensitivity to motion, muscle, and eye movement induced artifacts and requirement to apply conductive gels or solutions.

Near—infrared spectroscopy (NIRS) is a method of optical brain imaging that records changes in hemodynamics at a depth of up to 4 cm from the scalp. Near—infrared light (760 nm – 850 nm) is emitted through the subject’s skull, while the local changes in intensity of light absorption and scattering are recorded by a detector. The measured light intensity can be converted into estimations of cerebral total hemoglobin (HbT) and differentiated into its factions: oxygenated (HbO) and deoxygenated (HbR) hemoglobin [14]. Being much more expensive than EEG, NIRS is more convenient for practical use in a BCI circuit. It does not require electrode gel and is less sensitive to artifacts from patient movements. In addition, brain activity classification can rely on several simultaneously measured quantities, including oxy—, deoxy—, and total hemoglobin concentrations. Only a few articles have been published on the use of NIRS—BCI after stroke [15–17].

Due to the limited availability of NIRS technology, open access labeled NIRS datasets are highly valuable for rehabilitation BCI developers, particularly for validating machine learning and artificial intelligence algorithms for classifying brain signals. Currently, there are several available open access NIRS [18,19] or NIRS+EEG [20,21] datasets collected from 24—30 healthy subjects and containing 1—3 recordings from each participant. Data from stroke patients may differ due to the brain damage, potential changes in cognitive and neuropsychological functions, and older age.

To the best of our knowledge, this is the first open access dataset containing NIRS recordings from stroke patients. The dataset comprises 15 participants, 237 individual motor imagery BCI sessions utilizing three different mental tasks, over 50 hours of NIRS recordings, and 5296 trials. Each patient completed 7–24 online BCI training sessions. On average, the dataset includes 353 trials and 3.3 hours of NIRS recording data per participant.

We believe the data recorded from real stroke patients in multiple BCI sessions will be of particular interest to teams designing NIRS BCI systems for stroke rehabilitation and groups studying brain activity corresponding to motor imagery. The number of sessions is typical for a single hospitalization and allows to estimate how well cross—session transfer learning algorithms would work in real practice. The recordings from various patients could be utilized to develop and evaluate algorithms for cross–subject classification of hemodynamic activity related to motor imagery. Furthermore, the data could aid in studying the patients’ hemodynamic response to motor imagery and the response changes throughout the whole rehabilitation course.

## Materials and Methods

### Participants

This study included fifteen patients admitted to the post–stroke rehabilitation department: 9 males and 6 females, all right–handed, 58.8 [49.4; 70.0] years old (median, 25% and 75% quartiles); time since stroke onset was 7.0 [2.0; 10.0] months; all patients had one–sided cortical lesions, 8 in the left and 7 in the right hemisphere; the upper extremity Fugl–Meyer Assessment (UE–FMA) score at baseline was 47,0 [35,0; 54,0]; the Action Research Arm Test (ARAT) score at baseline was 35.0 [10.0; 44.0]. For more details refer to the Supplementary Information (see Table 1). All participants were informed about the experimental procedure and gave written consent prior to the experiment. This study was conducted according to the Helsinki declaration and was approved by the Local Ethics Committee of the Research Center of Neurology (approval number: No. 5– 4/22 dated June 1, 2022). The patients’ data were anonymized and depersonalized according to the local laws.

**Table 1.**
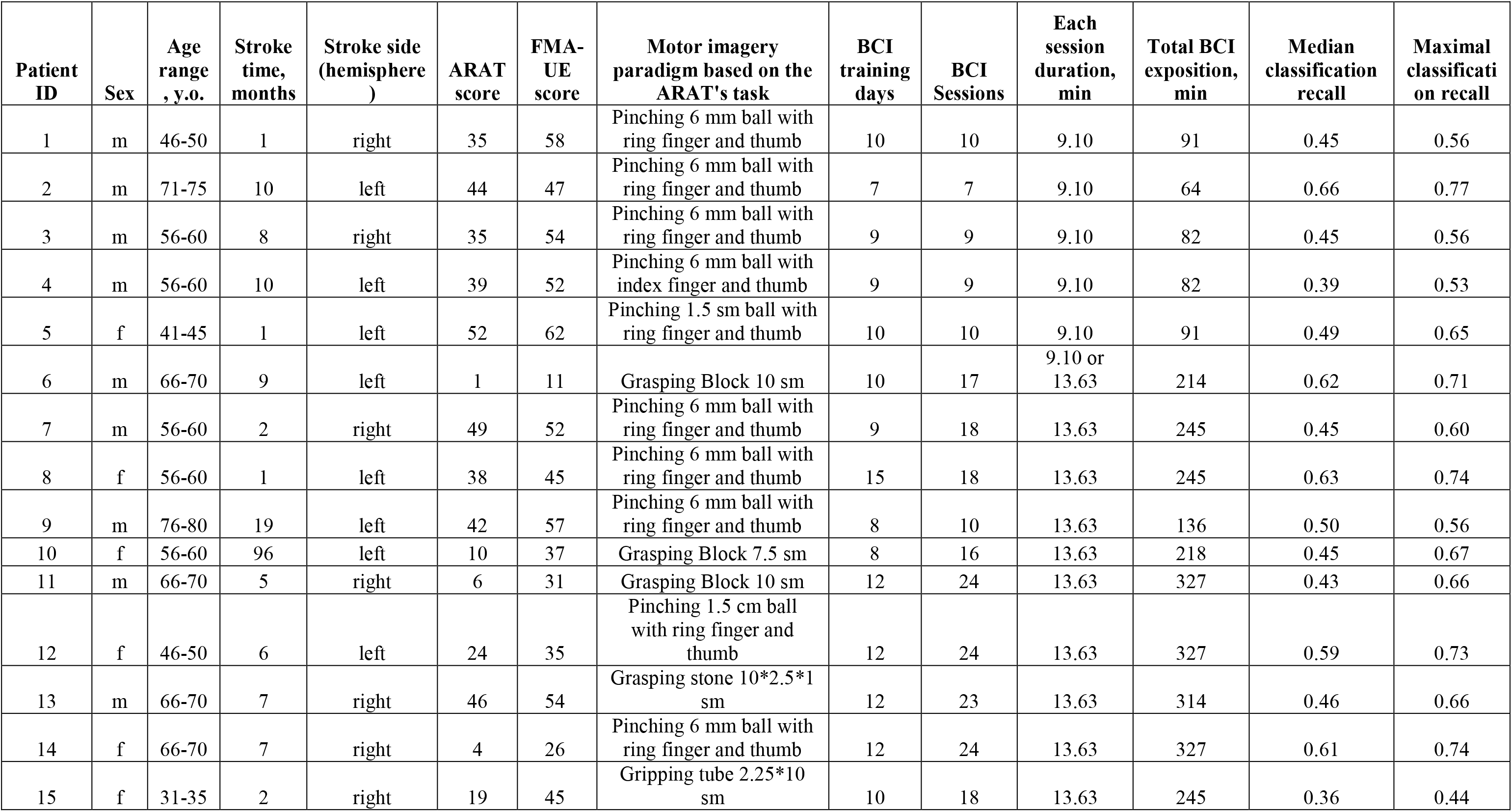
Subjects characteristics and classification accuracy.

### Experimental paradigm

The physical therapist individually selected the movement type for imaging for each patient. The selected movement was the most challenging among those included in the ARAT test (refer to Table 1). Prior to each training session, the physical therapist asked the patient to perform or attempt the target movement several times until they confirmed their readiness to mentally reproduce this movement – this is known as the priming step. If the target movement involved manipulating any ARAT subject (e.g. ball, wood block, or tube), it was provided to the patient during priming.

The patient, wearing a NIRS cap, sat in an armchair in front of a computer monitor with their hands resting on the armrests or the table. The screen displayed a black background with a fixation circle and three gray arrows in the center. The arrows corresponded to the tasks the patient was instructed to perform: the upper arrow indicated relaxation, while the left and right arrows corresponded to imagined movement of patient’s left and right hand, respectively. Changing the arrow color to blue served as a cue to prepare and changing the arrow color to green signaled to start performing the corresponding task. Correct classification was indicated by a green and enlarged circle, while an incorrect classification was indicated by a smaller circle. No feedback was provided when the patient had to relax or prepare. Figure 1 shows a typical session structure (<u>Figure 1</u>).

**Figure 1.**
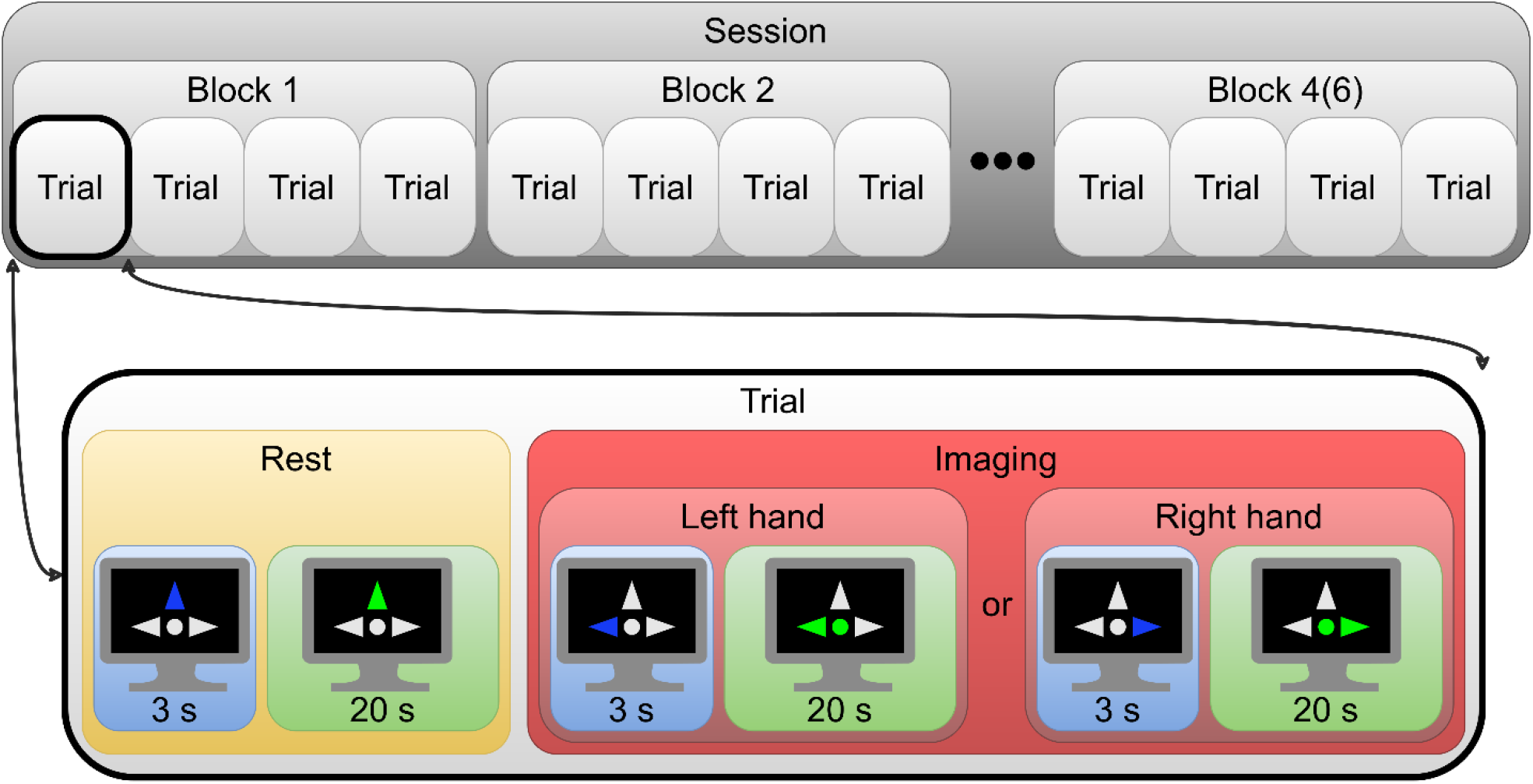
Typical session structure.

One experimental day with one patient consisted of one or two sessions. The study lasted from 7 to 15 days, with each patient participating in 7 to 24 sessions, totaling 237 sessions. A session comprised of 4 or 6 blocks and lasted 9 or 14 minutes. Each block included 4 trials: 2 right–hand movement imagining and 2 left–hand movement imagining, presented randomly. A single trial consisted of a 17–second relaxation phase followed by a 17–second movement imagery phase. During each phase, participants were given 2 seconds to prepare and 15 seconds to perform the corresponding task. Both movement imagery and relaxation were classified with overlapping epochs of 1 second and an epoch shift of 250 milliseconds. During the movement imagery, the feedback was updated based on the classification results.

### Data Acquisition

The data were acquired using a NIRScout (NIRx Medizintechnik GmbH, Berlin, Germany) with 8 detectors and 16 light sources at wavelengths of 760 and 850 nm. The sampling rate was 15.6 Hz. Fourteen sources and eight detectors were placed at a distance of about 3 cm from each other above the motor areas. Figure 2 shows the locations of all sources, detectors, and channels. The sources were positioned at F3, FC5, FC1, C3, CP5, CP1, P3, F4, FC2, FC6, C4, CP2, CP6, P4. The detectors were positioned at FC3, C5, C1, CP3, FC4, C2, C6, CP4. A total of 28 source–detector pairs were chosen to create the NIRS channels that were recorded.

**Figure 2.**
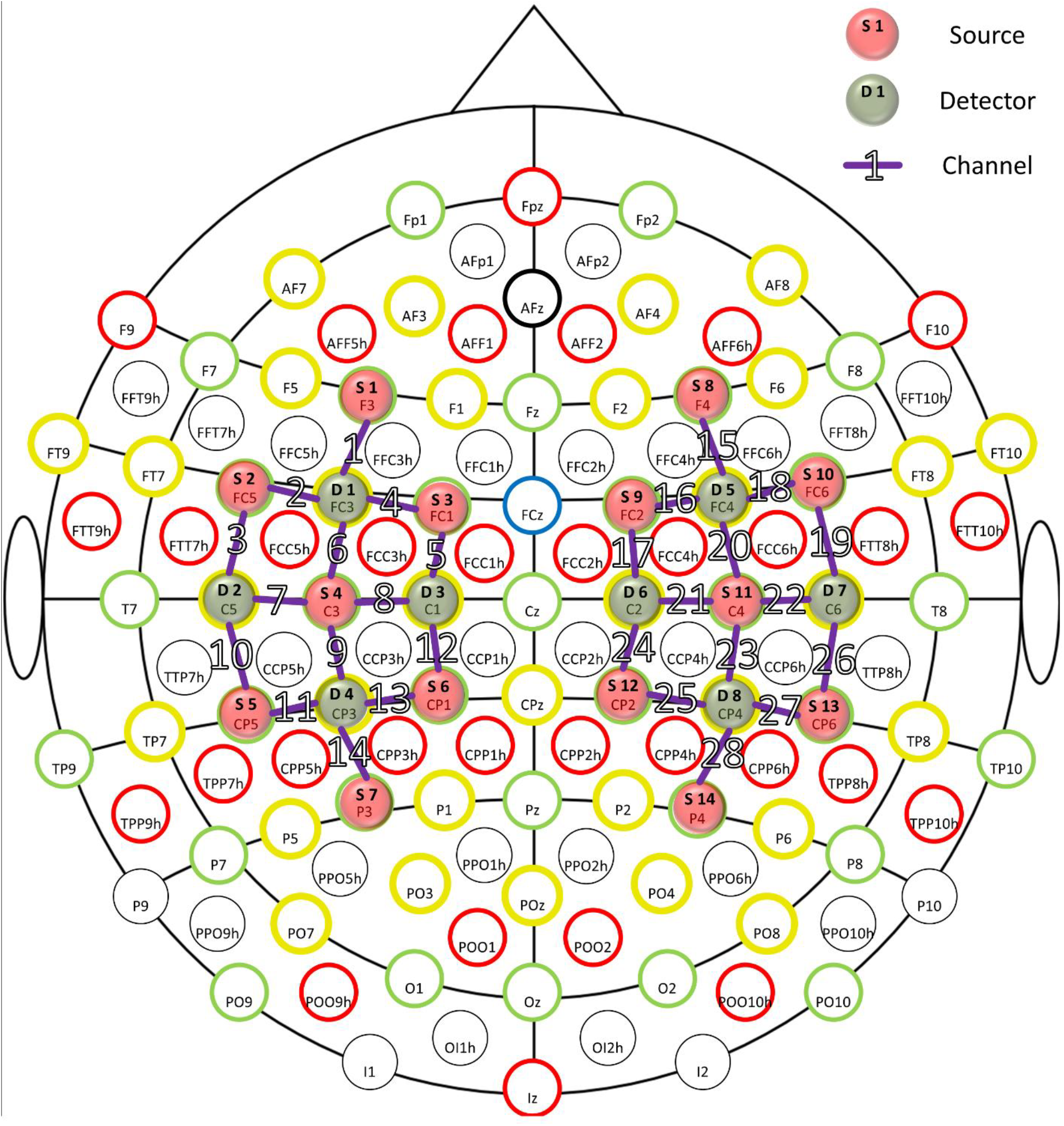
Positions of all sources, detectors, and channels. Red and green circles indicate the sources and the detectors respectively, purple lines indicate the channels.

### Online Data Processing

MATLAB R2019b (MathWorks, Natick, USA) was used for all data processing. The raw NIRS data were converted to oxy– and deoxyhemoglobin relative concentrations (HbO and HbR, respectively) using the modified Beer– Lambert law. The classification was performed in two steps: first, the classifier determined whether the epoch was related to relaxation or motor imagery. Next, if motor imagery was recognized, the classifier determined which hand movement was imagined. For the first classification step, the data were band–pass filtered (2nd order Chebyshev filter) with 0.022 Hz and 0.039 Hz cutoff frequencies. These frequencies were chosen to eliminate slow, high–amplitude signal trends and minimize phase shifts on the fundamental frequency (1/34 = 0.029 Hz) [22]. Shrinkage regularization was used to avoid the adverse effects of multicollinearity. For the second classification step, the data were high–pass filtered (1st order Chebyshev filter) with 0.005 Hz cutoff frequency. Filtered relative concentrations (HbO and HbR) were chosen as features. Linear discriminant analysis was used to classify the data in both steps. The training sample consisted of all previous blocks from the current and past sessions of the patient.

## Data Records

The data were presented in MATLAB format. Each filename includes subject ID, day and session number. The file contains a tabulated configuration of the channels (source–detector pairs in the 10–10 system), the raw light intensity data for each channel on both wavelengths, HbO and HbR concentrations in mmol/l, labels of mental tasks for each time point, and a confusion matrix of online classification.

## Technical Validation

The grand median recall for online classification of all subjects was 46.0 [44.7; 60.3]%. All subjects exceeded the random classification level. Classification performance varied significantly between patients and between sessions for most subjects (Figure 3).

**Figure 3.**
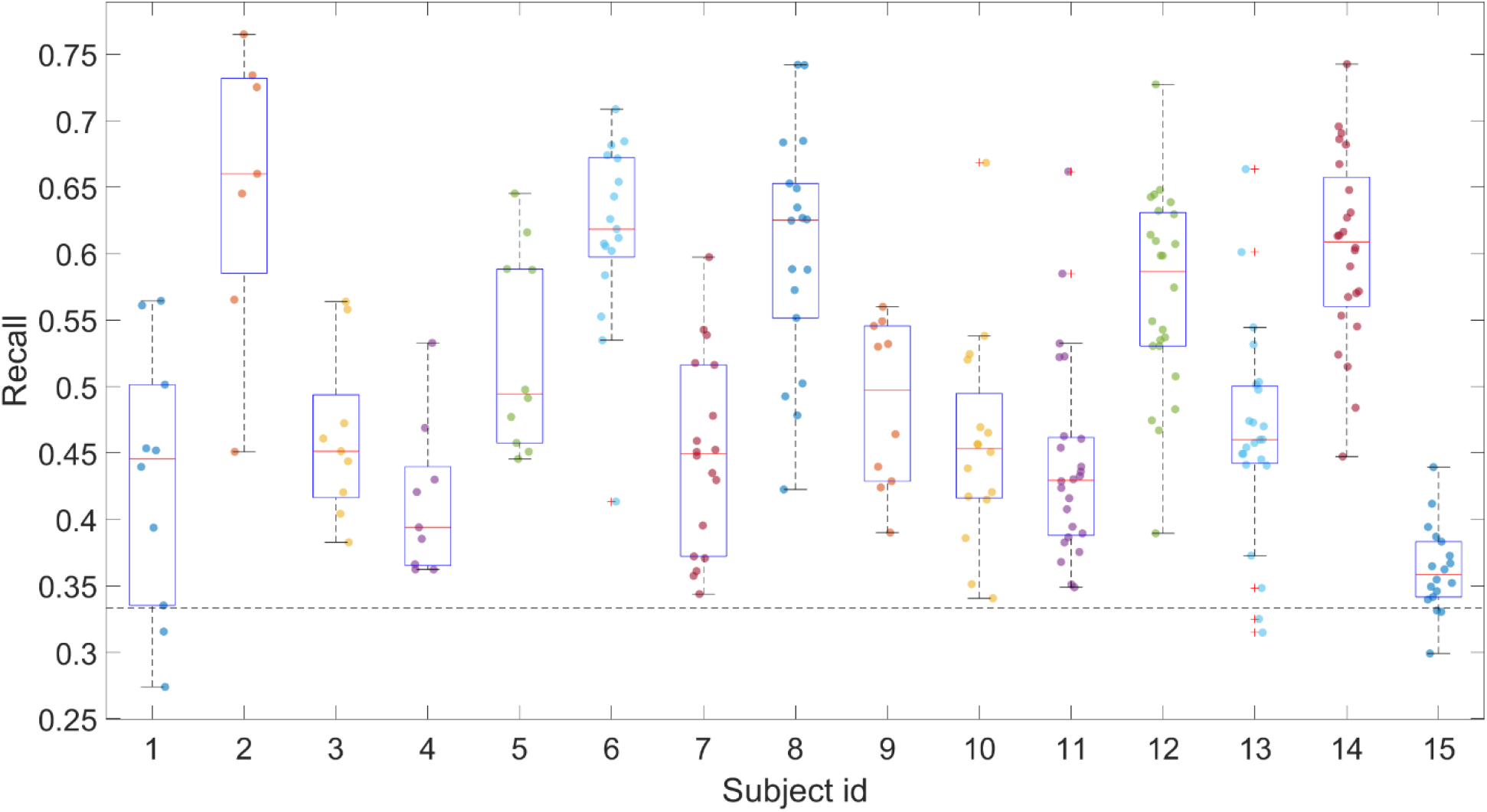
Online classification performance of all subjects. Dots indicate sessions, red lines indicate medians for each subject, boxes indicate 25% and 75% quartiles, dash line indicates random chance level.

The median within–subject range of session recall was 29.5 [18.6; 31.8]%. To plot the responses during left or right imaging, the data were zero phase band– pass filtered (4nd order Chebyshev filter) with 0.005 Hz and 0.09 Hz cutoff frequencies, and each response was baselined by subtracting the average value of the last 10 seconds before the task began. Averaging HbO and HbR responses separately for patients with left– and right–lesioned hemisphere shows that the response is present to the imaging of both hands in both hemispheres (<u>Figure 4</u>). At the same time, the response is similar for the imaging of both the paretic and intact hand in the intact hemisphere. However, there is an asymmetry of the response in the affected hemisphere, with a greater response to the imaging of the paretic hand. We suppose this asymmetry of hemodynamic response is due to a greater interhemispheric inhibitory drive from the intact hemisphere to the lesioned one [23,24].

**Figure 4.**
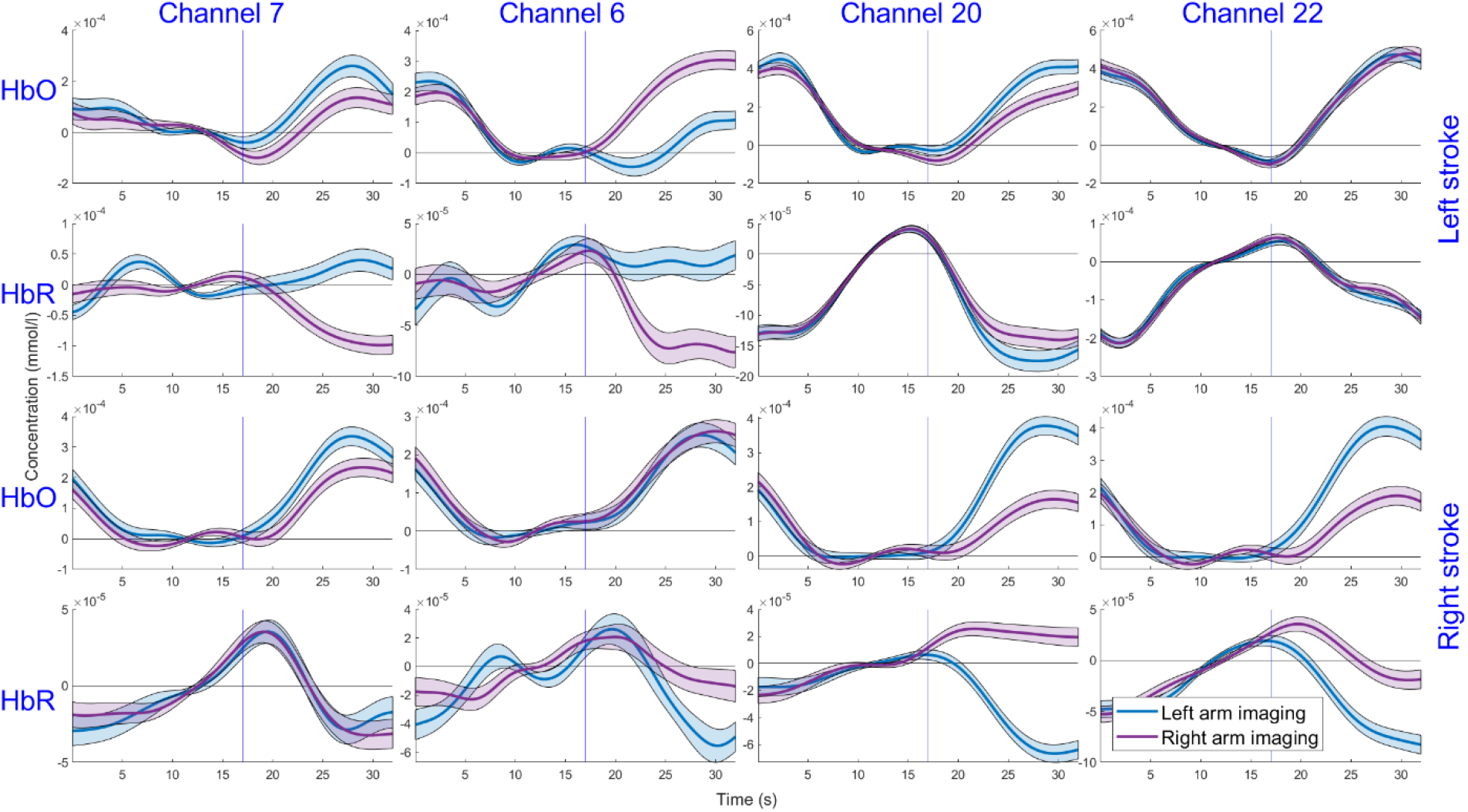
Hemodynamic responses during rest and imaging states at channels 6 and 7 from the left hemisphere and symmetric channels 20 and 22 from the right hemisphere for patients with left– and right–side strokes. The blue and red lines indicate left– and right–hand imaging. Semi–transparent areas show standard error and blue vertical line indicates movement imaging start.

## Usage Notes

The dataset is freely available at the Research Center of Neurology server at https://neurology.ru/dataset_nirs.rar.

This dataset is licensed under the Creative Commons Attribution-NonCommercial-NoDerivatives (CC BY-NC-ND).

One of the main disadvantages of the dataset is its unbalanced design (4 or 6 blocks per session and a different number of sessions for patients). Unfortunately, this is how real rehabilitation procedures look like: for various reasons patients stop participating in the experiment. If one wants to analyze balanced data, one can simply discard the extra blocks and sessions, the experiment design allows this to be done.

## Data Availability

All data produced in the present work are contained in the manuscript or available online at https://neurology.ru/dataset_nirs.rar

## Author contributions

M.R.I. — the BCI software development, the dataset preparation and technical validation, creating the illustrations, writing the manuscript draft

O.A.M. — development of the data collection protocol, writing the manuscript draft

R.Kh.L. — development of the study protocol and methods, subject screening

E.S.I., A.N.Ch. — subject screening, conducting the BCI trainings

N.A.S., P.M.A., P.D.B. — developing the methods for dataset preparation, justification of the study concept

All authors participated in editing the draft of the manuscript and its final version approval.

